# Explainable Machine Learning Models for Alzheimer’s Diagnosis Using Routine and Low-Cost Clinical Data

**DOI:** 10.64898/2026.07.10.26357720

**Authors:** Daniele De Carli, Alberto Sudati, Fabio Dercole

**Affiliations:** Politecnico di Milano; Department of Electronics, Information and Bioengineering Politecnico di Milano

**Keywords:** Neural Network, LightGBM, Alzheimer’s disease, Explainability

## Abstract

Emerging as a significant global health challenge, Alzheimer’s Disease (AD) is a progressive neurodegenerative disorder that causes memory loss and cognitive decline. Despite the ever-increasing waiting time for a specialist diagnosis, the need for a cost-effective and fast diagnostic technique is evident. This study explores the development of an explainable deep learning model to diagnose AD using only routine and low-cost clinical data, including demographic information, patient history, and results of neuropsychological tests (limited to those that can be automatically acquired). The analysis was carried out using a dataset provided by the National Alzheimer’s Coordinating Center, comprising 167,364 observations and 1,024 features. The findings demonstrate diagnostic performance comparable, and slightly superior, to that of clinicians when evaluated under similar informative constraints. This study introduces two classification models to discriminate whether the presumptive etiological cause of cognitive impairment is Alzheimer’s disease. The deep neural network achieved an accuracy of 90% with an area under the receiver operating characteristic curve (ROC-AUC) of 0.96, whereas the Light Gradient Boosting Machine reached the same accuracy with a ROC-AUC of 0.97.

## 1 Introduction

Dementia affects more than 57 million people in 2021 and this number is expected to reach 152 million by 2050^1,2^. Alzheimer’s disease (AD)^3^ is the most common cause of dementia worldwide^4^. Despite decades of research, the disease remains incurable, and early diagnosis is one of the most effective strategies currently available to slow cognitive decline and improve patient outcomes^5^. However, existing diagnostic pathways often rely on expensive^6,7^, invasive, or time-consuming^8^ methods, such as brain imaging and cerebrospinal fluid (CSF) analysis, which limits their accessibility and scalability, particularly in low-resource settings.

In this context, artificial intelligence (AI) and deep learning have emerged as promising tools to enhance early detection by leveraging large-scale clinical datasets^9^. Recent studies have focused on complex, multimodal architectures combining imaging, biomarkers, and cognitive features to achieve high diagnostic accuracy^10,11^. Although these methods show excellent performance, their real-world applicability is constrained by the cost and availability of the required data sources.

Despite the growing body of research on AI-based diagnosis of Alzheimer’s disease, existing approaches present important trade-offs between predictive performance, interpretability, and real-world applicability. For instance, lightweight models based on routine clinical data have demonstrated moderate performance, often relying on aggressive feature selection strategies that limit the representation of the patient’s clinical profile^12^. Conversely, more recent explainable AI frameworks operating on high-dimensional feature spaces have achieved strong predictive performance, but typically without constraints on data accessibility or acquisition cost^13^.

In parallel, studies comparing clinical diagnoses with neuropathological confirmation have highlighted the intrinsic limitations of diagnostic accuracy in real-world settings, particularly when advanced biomarkers are not available^14^. These observations underscore the need for approaches that balance predictive accuracy, interpretability, and scalability, while remaining grounded in clinically accessible data.

This study proposes an alternative approach that focuses on simplicity, accessibility, and explainability. We explore the potential of machine learning models trained exclusively on routinely available non-invasive clinical data such as demographic information, medical history, and standardized cognitive test scores selected from the National Alzheimer’s Coordinating Center (NACC) Uniform Data Set. These variables are inexpensive to acquire and widely used in clinical practice, making them ideal candidates for large-scale, low-cost screening strategies.

Several cognitive assessments commonly used in the diagnosis of Alzheimer’s disease have been the subject of successful automation efforts, as documented in the literature. Examples include the Story Recall Task (`CRAFTURS`)^15^, Trail Making Test (`TRAILA`, `TRAILALI`), Mini-Mental State Examination (`NACCMMSE`)^16^, and the Wechsler Adult Intelligence Scale (`WAIS`)^17,18^. Another cognitive test, the Montreal Cognitive Assessment (MoCA) (`NACCMOCA`), now offers an online self-testing tool to obtain the MoCA score. More sophisticated assessments, such as the Rey Complex Figure Test^19,20^, have been successfully automated using convolutional neural networks, illustrating the feasibility of digitizing even highly intricate cognitive evaluations. Another study^21^ explores human-robot interaction (HRI) for automatic scoring of a revised version of the MoCA test, using visual-spatial and speech recognition.

Drawing on evidence from the literature, we conclude that artificial intelligence can enable the creation of a rapid, cost-effective, and scalable platform for screening and preliminary diagnosis of cognitive decline. By allowing patients to complete a battery of automated cognitive assessments, either independently or with the help of a caregiver, where necessary, the resulting data can be seamlessly integrated into an AI-driven system that provides accurate and interpretable evaluations. Such an approach is particularly valuable for expanding access to early detection in developing countries^22^, where traditional diagnostic infrastructure is often limited while the burden of neurodegenerative diseases increases^23^. This scalable and accessible solution has the potential to overcome critical barriers in global healthcare delivery, facilitating timely intervention and reducing disparities in disease management.

The goal of this work is twofold: first, to develop machine learning models, in particular using Deep Neural Network (DNN), and Light Gradient Boosting Machine (Abbreviations: LightGBM or LGBM) a high-performance Gradient Boosting Decision Trees (GBDT) farmework, capable of accurately distinguishing between AD and non-AD cognitive states; second, to ensure that these models remain interpretable and clinically transparent through SHAP based explanation techniques (SHapley Additive exPlanations)^24^ (the importance and expected results of Machine Learning explainability is discussed in section 2.8).

Although the study demonstrates promising results in differentiating Alzheimer’s disease (AD) from non-AD cognitive states using routine and low-cost clinical data, several limitations warrant consideration. In particular, the work by Aghdam et al.^25^ highlights the challenges in distinguishing between stable mild cognitive impairment (sMCI) and progressive cognitive impairment (pMCI). Since our classification task encompasses all stages of AD versus all non-AD states, cases with intermediate or uncertain cognitive profiles, such as MCI, may fall near the decision boundary, potentially reducing model confidence and accuracy. This limitation underscores the intrinsic difficulty of classifying early or borderline cognitive impairments using routine and low-cost clincal data alone, without the additional resolution provided by neuroimaging or molecular biomarkers.

Looking forward, an important direction for this project involves developing a multiclass classification framework capable of distinguishing among distinct stages of cognitive decline, from early impairment to advanced Alzheimer’s disease, using only tabular clinical and neuropsychological data. This vision aligns with complementary future efforts to leverage longitudinal disease trajectories through temporal deep learning and survival analysis, ultimately enabling more precise, non-invasive, and personalized patient stratification in clinical and research settings.

The article begins by describing the dataset used in this study (Sect. 2.1). The methodological framework is then outlined, covering feature engineering, machine learning (ML) techniques, hyperparameter tuning, overfitting control, and ML explainability via SHAP (Sects. 2.2 -2.8). The results are presented and discussed in Sect. 3, including a comparison with existing tools in Sect. 3.4. The article concludes with final remarks and information on data and code availability (Sect. 4).

## 2 Methods

A comprehensive overview of the experimental workflow, encompassing data acquisition, model optimization, and explainability analysis, is synthesized in Figure 1.

**Figure 1:**
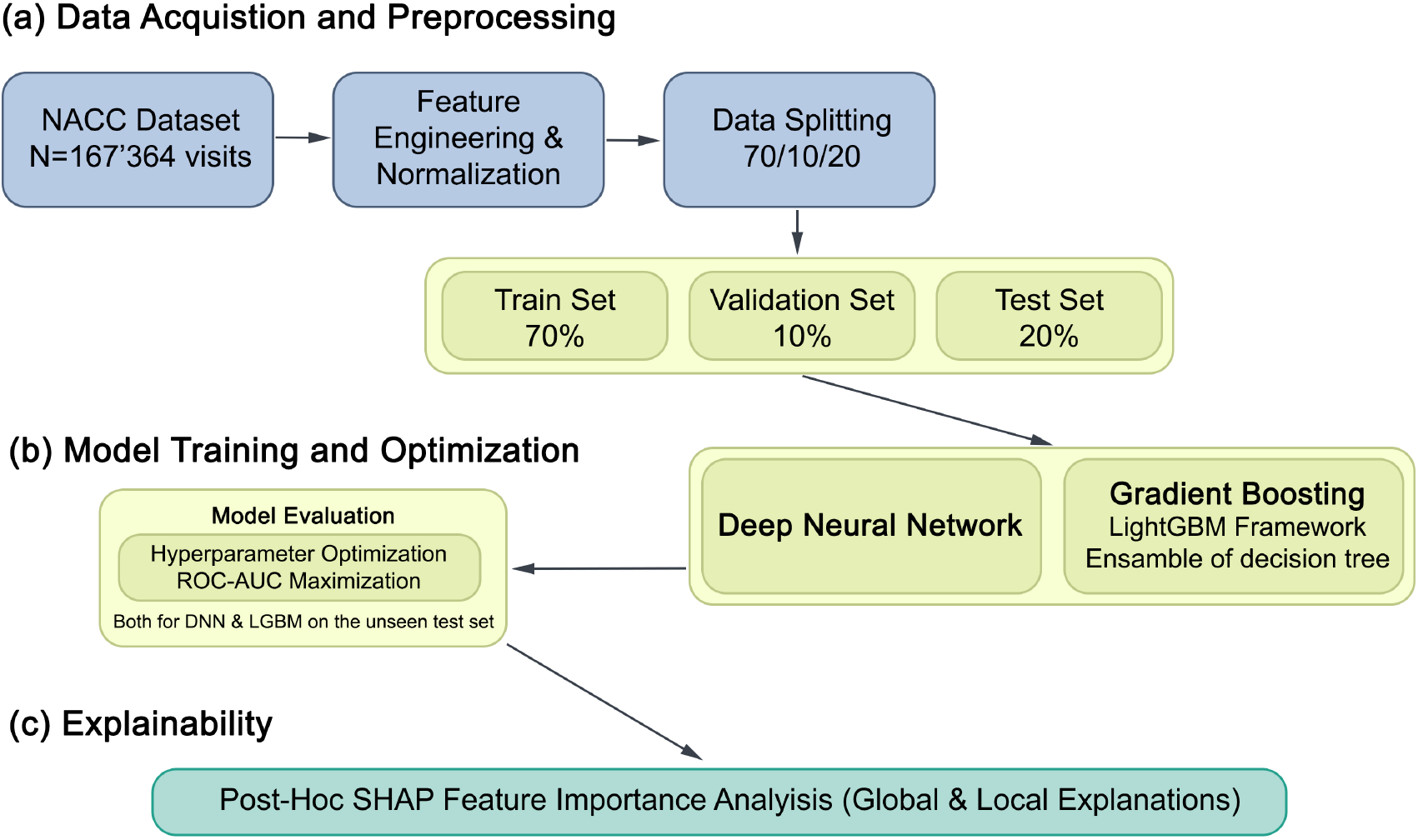
The study progressed through three phases: **(a)** curation and preprocessing of the NACC dataset (*N* = 167, 364 visits); **(b)** training and hyperparameter optimization of Deep Neural Network (DNN) and LightGBM architectures using the Optuna framework; and **(c)** SHAP-based feature importance analysis (on the independent test set) to ensure global and local interpretability of the predictive outcomes.

### 2.1 NACC Dataset

Data for this study were obtained from the National Alzheimer’s Coordinating Center (NACC) database through a formal research data request, in accordance with NACC data access policies. Analyses focused on the Uniform Data Set (UDS), a core component of the NACC repository that comprises systematically collected clinical visit data acquired under harmonized protocols across participating centers. The UDS includes demographic information, medical history, cognitive status, and standardized neuropsychological assessments. A detailed description of the dataset is provided in^26^.

At the time of data extraction, the dataset incorporated contributions from 186 Alzheimer’s Disease Research Centers (ADRCs), identified through the NACCADC variable, reflecting the diversity and evolution of participating centers over time. The time range covers UDS visits conducted between September 2005 and June 2024. Each participant may contribute multiple visits over time, enabling longitudinal tracking of cognitive and clinical changes.

### 2.2 Feature Selection

The selection of features was guided by the overarching goal of the project: to develop a predictive model based on data that are easily obtainable and routinely available in clinical settings. To accomplish this task, the selected features encompass all available variables related to the individual’s medical history, demographic information, and the results of cognitive evaluations. Table 1 contains the features used in this study.

**Table 1:**
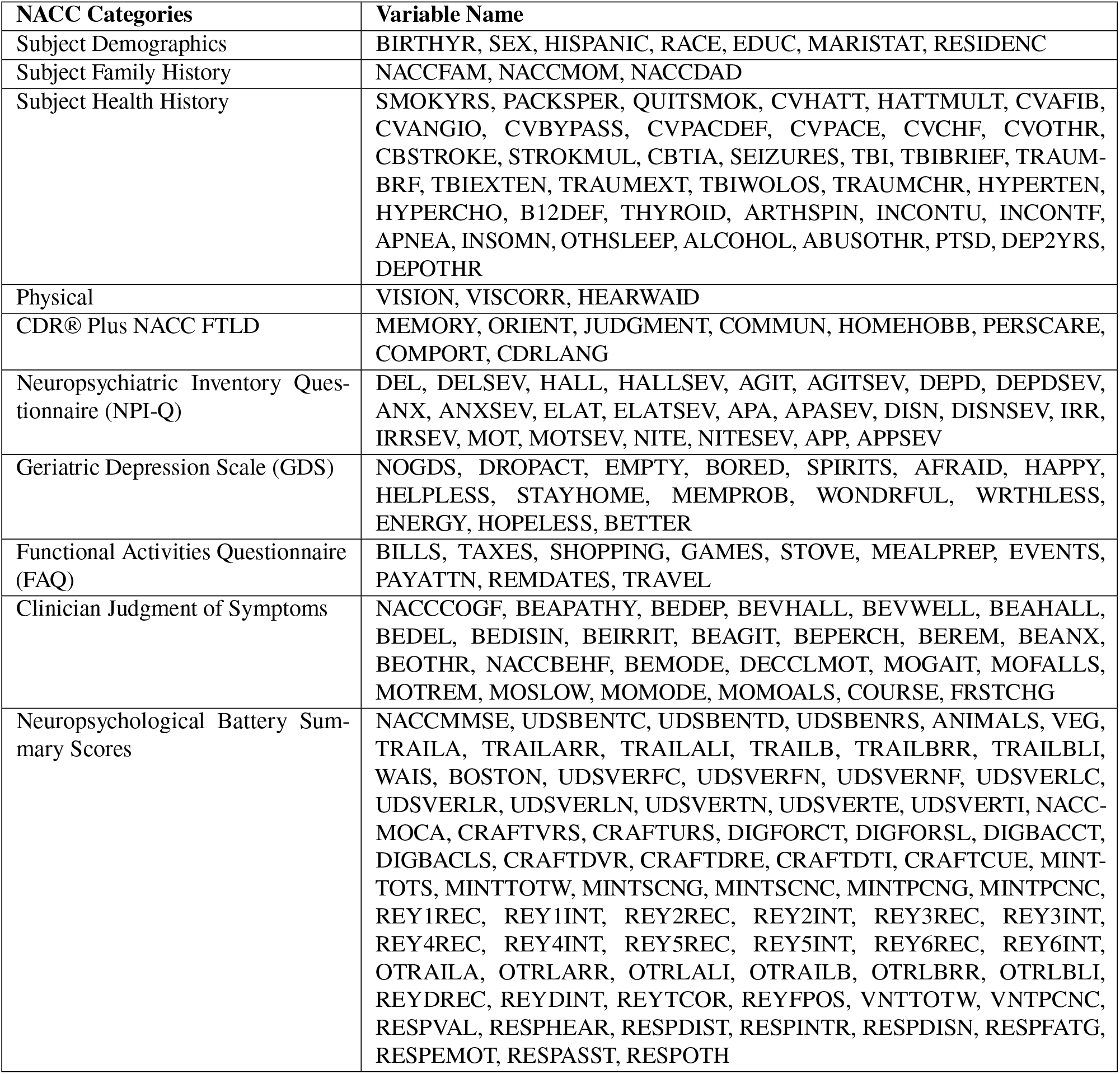
Features Selected in the first stage of the study.

**Table 2:** Hyperparameter search space for DNN.

| Hyperparameter | Range / Options | Description |
| --- | --- | --- |
| threshold | Float in [0.900, 0.999] | Correlation-based feature elimination. |
| n_layers | Integer in [1, 6] | Number of hidden layers. |
| units_l{i} | Integer in [16, 512] (step=16) | Neurons in the $i^{\text{th}}$ layer. |
| dropout_rate_l{i} | Float in [0.0, 0.4] | Dropout rate after the $i^{\text{th}}$ layer. |
| learning_rate | Float in [$10^{-5}$, $10^{-2}$] (log-scale) | Learning rate for the Adam optimizer. |
| batch_size | Categorical: {8, 16, 32, 64, 128} | Batch size used during training. |

**Table 3:**
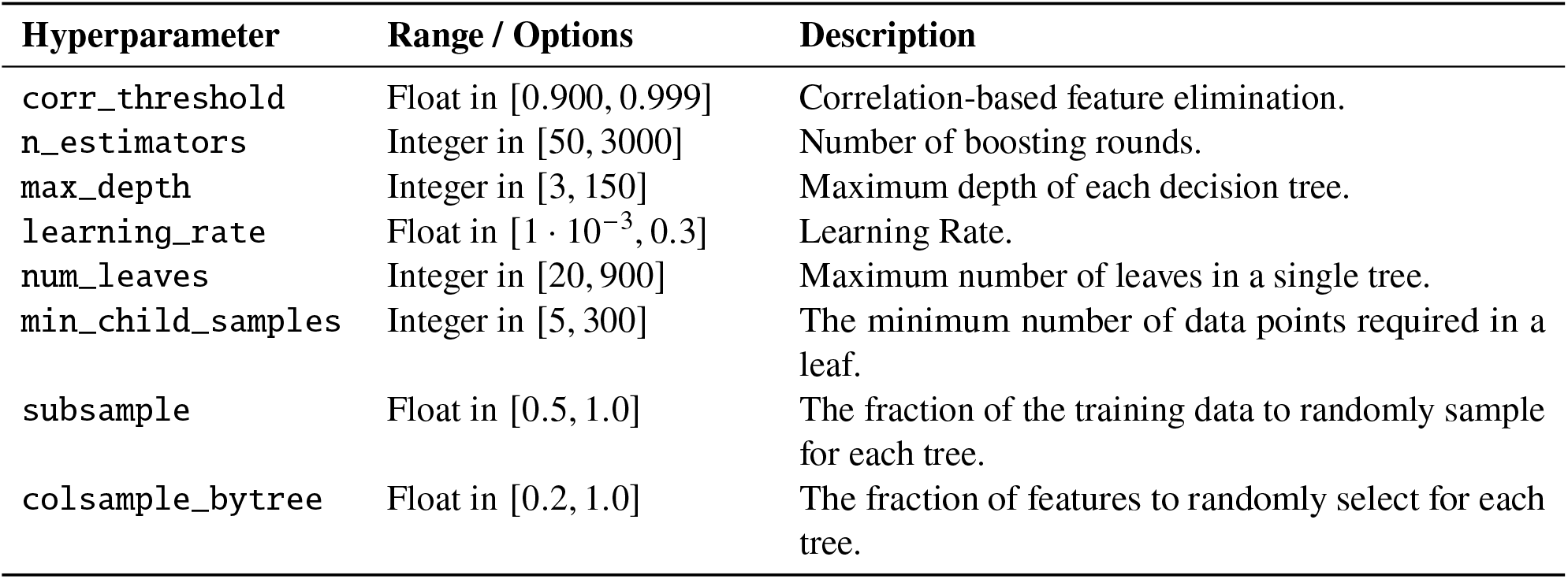
yHyperparameter search space for LightGBM.

| Hyperparameter | Range / Options | Description |
| --- | --- | --- |
| corr_threshold | Float in [0.900, 0.999] | Correlation-based feature elimination. |
| n_estimators | Integer in [50, 3000] | Number of boosting rounds. |
| max_depth | Integer in [3, 150] | Maximum depth of each decision tree. |
| learning_rate | Float in [$1 \cdot 10^{-3}$, 0.3] | Learning Rate. |
| num_leaves | Integer in [20, 900] | Maximum number of leaves in a single tree. |
| min_child_samples | Integer in [5, 300] | The minimum number of data points required in a leaf. |
| subsample | Float in [0.5, 1.0] | The fraction of the training data to randomly sample for each tree. |
| colsample_bytree | Float in [0.2, 1.0] | The fraction of features to randomly select for each tree. |

**Table 4:** Optimal hyperparameters for the best-performing neural network model.

| Hyperparameter | Optimal Value |
| --- | --- |
| Number of hidden layers ($n_{\text{layers}}$) | 3 |
| Units in Layer 1 | 256 |
| Units in Layer 2 | 192 |
| Units in Layer 3 | 64 |
| Dropout rate Layer 1 | 0.02251 |
| Dropout rate Layer 2 | 0.10849 |
| Dropout rate Layer 3 | 0.15882 |
| Learning rate | $2.71250 \times 10^{-4}$ |
| Batch size | 8 |
| Correlation threshold | 0.99002 |

**Table 5:** Optimal hyperparameters for the best-performing GBDT model.

| Hyperparameter | Optimal Value |
| --- | --- |
| corr_threshold | 0.97037 |
| n_estimators | 2767 |
| max_depth | 83 |
| learning_rate | 0.00822 |
| num_leaves | 72 |
| min_child_samples | 90 |
| subsample | 0.52338 |
| colsample_bytree | 0.24950 |

**Table 6:** Performance comparison between Deep Neural Network (DNN) and Light Gradient Boosting Machine (LGBM) models.

| Model | Accuracy | Sensitivity | Specificity | Precision | F1-score | AUC |
| --- | --- | --- | --- | --- | --- | --- |
| DNN | 0.8953 | 0.8770 | 0.9052 | 0.8326 | 0.8542 | 0.9606 |
| LGBM | 0.9024 | 0.8792 | 0.9148 | 0.8473 | 0.8630 | 0.9674 |

*Abbreviations:* AUC, Area Under the ROC Curve.

It is important to note that although these features were considered during the exploratory and preparatory phases, not all of them were ultimately used in model training. The rationale behind this exclusion, along with the criteria adopted for feature selection in the final models, will be discussed in Section 2.4.

### 2.3 Target Variable

The target variable NACCALZD is a categorical longitudinal indicator derived from the UDS, capturing the presumed etiologic diagnosis of cognitive impairment across repeated clinical assessments, with Alzheimer’s disease as the etiologic outcome of interest. The variable NACCALZD is assigned to each patient visit. It was originally encoded as follows:

- 0: Cognitive impairment is present, but it is not attributed to AD.
- 1: Any cognitive impairment (dementia, MCI, or impaired, not MCI) attributed to AD.
- 8: No cognitive impairment (i.e., cognitively normal).

To enable binary classification, distinguishing individuals affected by Alzheimer’s disease from those not affected, the original value 8 was recoded to 0. This transformation consolidates all non-AD cases into a single class and reflects the objective of the modeling: to classify subjects as AD positive (AD = 1) or AD negative (AD = 0). This binarization aligns with the study’s goal of training a predictive model capable of reliably identifying Alzheimer’s related cognitive impairment in contrast to all other cognitive states.

### 2.4 Normalization and Feature Engineering

Pre-processing steps targeting codified categorical features were applied prior to normalization. Specifically, variables encoded using discrete categorical schemes in the dataset were processed to handle missing values and special administrative codes, whereas continuous variables (e.g., age or dates) were excluded from this procedure.

All categorical variables underwent a rule-based recoding procedure to standardize missing values and administrative flag codes. Missing values originally encoded as −4 were uniformly mapped to −1. In addition, extended categorical codifications were systematically remapped to distinct negative values to preserve their semantic role as non-physiological administrative indicators while preventing distortion of the normalization range.

This step was crucial in mitigating the detrimental effects of extreme outlier values on the normalization process. In Min-Max scaling, the presence of such outliers can substantially increase the range of features, anchoring the maximum value to a distant point in the distribution. As a consequence, the normalized values of the remaining observations are disproportionately compressed toward the lower bound of the [0, 1] interval, thereby reducing the effective dynamic range of the feature. This compression can hinder the model’s capacity to capture and exploit meaningful variability within the data, ultimately impairing the learning process. The applied adjustment thus ensures a more balanced feature scaling, preserving the discriminative structure of the input space.

Following this step, all feature columns, both processed categorical variables and continuous features, were scaled using Min–Max normalization to map values into the [0, 1] interval:

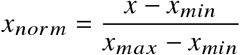

Where: *x* is the original value; *x*_*min*_ is the minimum value in the feature column; *x*_*max*_ is the maximum value in the feature column; *x*_*norm*_ is the normalized value.

Subsequently, the dataset underwent two additional pre-processing steps:

- **Low-Variance Feature Removal**: Features with a variance below 2 · 10^−6^ were eliminated, as they provided negligible discriminative power.
- **Correlation-Based Feature Elimination**: Highly correlated features (the correlation coefficient larger than a threshold *γ* found by optimizing the model’s hyperparameter, see subsection 2.7) were removed, reducing multicollinearity and the risk of overfitting the model. In particular, between two features with correlation above threshold, the one with the lower variance value was removed.

### 2.5 Dataset Splitting

The cleaned and normalized dataset was indexed by unique patient identifier, which were used to perform patient-level random shuffling before partitioning the data into three mutually exclusive subsets: 70% for training, 10% for validation, and 20% for testing.

Data splitting was performed by filtering all observations associated with each patient ID, ensuring that no subject contributed samples to more than one subset. In electroencephalography (EEG) studies, this strategy is critical to avoid data leakage, where samples from the same patient appearing in multiple sets could lead to artificially inflated model performance due to intra-subject correlations^27^. Without this precaution, the model could learn patient-specific patterns during training and unfairly leverage them during evaluation, violating the assumption of independent and identically distributed (*i.i.d*.) data and compromising the generalization capability of the neural network. Although this work does not use EEG, this precaution is nonetheless used.

### 2.6 Implemented Machine Learning Models

#### 2.6.1 Deep Neural Network (DNN) Architectures

The implemented model is a feedforward neural network (Multi-Layer Perceptron, MLP), constructed using the Keras Sequential API. The input dimension corresponds to the number of features in the pre-processed dataset. The network ends with a single neuron that uses a sigmoid activation function to support the binary classification between AD and non-AD individuals. The general architecture is defined as follows:

- **Input Layer**: Takes in the normalized feature vector **x** ∈ ℝ ^204^.
- **Hidden Layers**: A variable number of hidden layers, ranging from 1 to 6 (selected via hyperparameter optimization). Each layer consists of:
  - A `Dense` layer with ReLU activation function: ϕ(*x*) = max(0, *x*)
  - An optional `Dropout` layer to improve generalization and prevent co-adaptation.
- **Output Layer**: A single neuron with sigmoid activation function 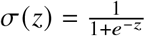, providing a score for the positive class (i.e., AD individual).

##### DNN Overfit control strategies

Several strategies were implemented to mitigate overfitting:

- **Layer-wise Dropout Regularization**: Dropout is applied independently to each hidden layer, and its rate is selected via hyperparameter tuning. Dropout helps prevent overreliance^28^ on specific neuron activations by randomly setting them at zero during training. The formal operation is:

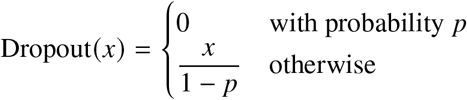
- **Early Stopping**: The model is trained for a maximum of 40 epochs, with early stopping implemented if the validation loss fails to improve for 10 consecutive epochs, restoring the best weights observed during training^29^.
- **LossRatioStopCallback**: An early stopping criterion that stops training if the ratio between validation and training loss exceeds a threshold (default: 2), indicating potential overfitting^30^:

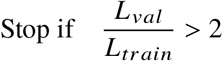

#### 2.6.2 LightGBM (LGBM) Architecture

In addition to the neural network model, we employed a Gradient Boosting Decision Tree (GBDT) approach implemented via the LightGBM framework. It is an efficient, distributed, and high-performance gradient boosting library that constructs decision trees in a leaf-wise manner with depth constraints, optimizing both computational speed and predictive accuracy. The model operates by sequentially combining multiple decision trees, where each tree partitions the feature space through a series of threshold-based splits on individual features, and each input sample is routed from the root to a leaf node that assigns a scalar score. The final prediction is obtained by aggregating the outputs of all trees and applying a logistic transformation to produce a probability for the positive class.

The model was trained on the preprocessed dataset, with the objective of binary classification between AD and non-AD individuals.

The general configuration is as follows:

- **Input Features**: Takes in the normalized feature vector **x** ∈ ℝ^192^. The correlation threshold used for feature elimination was optimized during model tuning, which results in an input dimension differing from that of the deep neural network.
- **Boosting Framework**: The LightGBM algorithm iteratively fits an ensemble of decision trees, where each subsequent tree corrects the residual errors of the previous ensemble. A leaf-wise growth strategy with a maximum depth constraint is employed, enabling the model to capture complex nonlinear relationships while controlling overfitting.
- **Regularization and Optimization**:
  - The parameters `subsample` and `colsample_bytree` introduce randomness in data and feature sampling, improving robustness.
  - Learning Rate controls the contribution of each tree to the ensemble, ensuring stable convergence.
- **Output Layer**: The final prediction is generated through a logistic transformation of the aggregated tree outputs, producing a probability score for the positive class (AD individual).

### 2.7 Hyperparameter Tuning

Hyperparameter optimization was performed to maximize the model’s ability to discriminate between classes during training. Specifically, the objective function was defined to minimize the complement of the maximum validation ROC AUC across epochs:

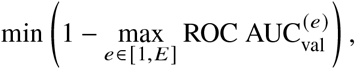

where ROC 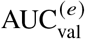 is the area under the receiver operating characteristic curve computed on the validation set at epoch *E*, summarizing the model’s overall ability to distinguish between positive and negative cases.

The optimization process was carried out using the Optuna Library^31^ and it was terminated after 106 trials for both models, once no substantial performance gains were observed. The training and hyperparameter search procedures were executed on Kaggle‘s cloud platform with an NVIDIA Tesla P100 GPU accelerator.

#### 2.7.1 DNN Hyperparameters Tuning

The following hyperparameters and the threshold for correlation-based feature elimination, see subsection 2.4, were included in the search space:

#### 2.7.2 LightGBM Hyperparameters Tuning

The following LightGBM hyperparameters and the threshold for correlation-based feature elimination subsection 2.4 are contained in the following table:

### 2.8 Explainability

The reliability and transparency of artificial intelligence models are essential requirements in clinical settings, where automated predictions must be interpretable and clinically justifiable. To this end, model explainability^32^ was assessed using an additive feature attribution framework based on Shapley values^24^, which originates from cooperative game theory. In this paradigm, each feature is interpreted as a “player” contributing to the final prediction, and its importance is quantified as the average marginal contribution to the model output across all possible feature coalitions (subsets of features considered jointly while others are treated as unknown or replaced by reference values).

The Shapley value of a given feature is computed by evaluating how the model prediction changes when that feature is added to different subsets of other features, then averaging this effect over the combinatorial space of all coalitions. Unlike global metrics that average across different data samples, this process occurs within a single instance, reflecting the feature’s marginal contribution while accounting for its complex interactions with other variables. This formulation enables a consistent decomposition of a prediction into feature-level attributions relative to a baseline output, ensuring local accuracy (the sum of attributions equals the prediction) and global consistency.

This explanation framework is particularly well suited to complex, non-linear models, as it relies solely on the input–output behavior of the trained model and does not require access to internal parameters, thereby allowing the analysis of black-box predictors.

#### Global and Local Explanations

Explainability was investigated at both the global and local levels. Global explanations aim to identify systematic patterns of feature influence across the entire dataset, providing insight into the overall decision-making strategy learned by the model, beyond individual predictions.

Given the heterogeneous nature of the adopted models, global explanations were computed using a model-specific SHAP implementation tailored to each architecture:

- For the optimized deep neural network (DNN), global feature importance was estimated using the `KernelExplainer`, which approximates Shapley values through a model-agnostic sampling strategy. Due to its computational cost, the analysis was performed on 250 randomly selected samples from the test subset.
- For the LightGBM model, global explanations were obtained using the `TreeExplainer`, which exploits the tree structure of gradient-boosted decision trees to compute exact Shapley values efficiently. This reduced computational complexity allowed the analysis to be conducted on the entire test subset.

The use of different SHAP implementations, KernelExplainer for the DNN and TreeExplainer for LGBM, does not invalidate the comparative analysis of feature importance. Both algorithms converge toward the same theoretical Shapley values (Global explenation results in Sect. 3.3, ensuring that the relative ranking and directional influence of features remain consistent and comparable across the two architectures.

In addition to global trends, the framework also provides **local explanations**, enabling the interpretation of individual predictions. At this level, the contribution of each feature is quantified in terms of how it shifts the model output toward or away from the predicted class for a specific patient, thus supporting patient-specific risk assessment and clinical interpretability.

## 3 Results & Discussion

The following sections present the optimal neural network architecture, its performance metrics, a comparison with existing literature, and interpretability analyses aimed at elucidating the model decision-making process to human experts.

### 3.1 Optimal ML Models Configurations

#### 3.1.1 Optimal DNN Hyperparameters

The architecture identified as optimal through hyperparameter optimization consisted of a multilayer feedforward perceptron with the following configuration:

#### 3.1.2 Optimal LGBM Hyperparameters

The best-performing architecture consisted of a Gradient Boosting Decision Tree model with the following configuration:

### 3.2 ML Model Test Performance

Two significant experiments have been conducted (detailed in subsection 2.6). For both DNN and LGBM models, detailed metrics are discussed to compare the models’ performance.

The LGBM model achieved its best performance in under five minutes of training, whereas the deep neural network required approximately 11 hours to reach its best-performing configuration. Note that these times reflect individual model training with hyperparameters selected during the preceding optimization process.

In terms of evaluation metrics, LGBM marginally outperformed DNN, achieving higher accuracy (90.24% vs. 89.53%), precision (87.92% vs. 87.70%) and AUC (0.9674 vs. 0.9606), indicating not only faster convergence, but also a modest improvement in performance across the independent test set.

The LGBM model demonstrated superior performance on tabular data, consistent with its known suitability for such datasets, benefitting from its native ability to handle categorical features and offering faster training with lower memory requirements compared to deep neural networks^33^.

### 3.3 Model Explanations and Feature Importance

To enhance model transparency and support clinical interpretability, we employed SHAP (SHapley Additive exPlanations) to quantify the contribution of each input feature to the model’s predictions. This allowed both a global assessment of the importance of the feature and localized patient-specific explanations. The following results highlight how different clinical variables influence the model decision-making process.

#### Global Importance of Features

The 20 most significant variables are reported in Figure 2, offering a graphical representation that provides a summarized global view of the average influence of each attribute in both the DNN and LGBM models. The differing x-axis scales between the two panels arise from the output space in which SHAP explanations are computed. In particular, SHAP values for the LGBM model are expressed in the log-odds space of the raw model output, leading to larger absolute magnitudes, whereas SHAP values for the DNN are computed in the probability space after the final activation function, resulting in comparatively smaller contribution values. As a consequence, SHAP magnitudes are not directly comparable across models and should be interpreted within each model as relative indicators of feature importance.

**Figure 2:**
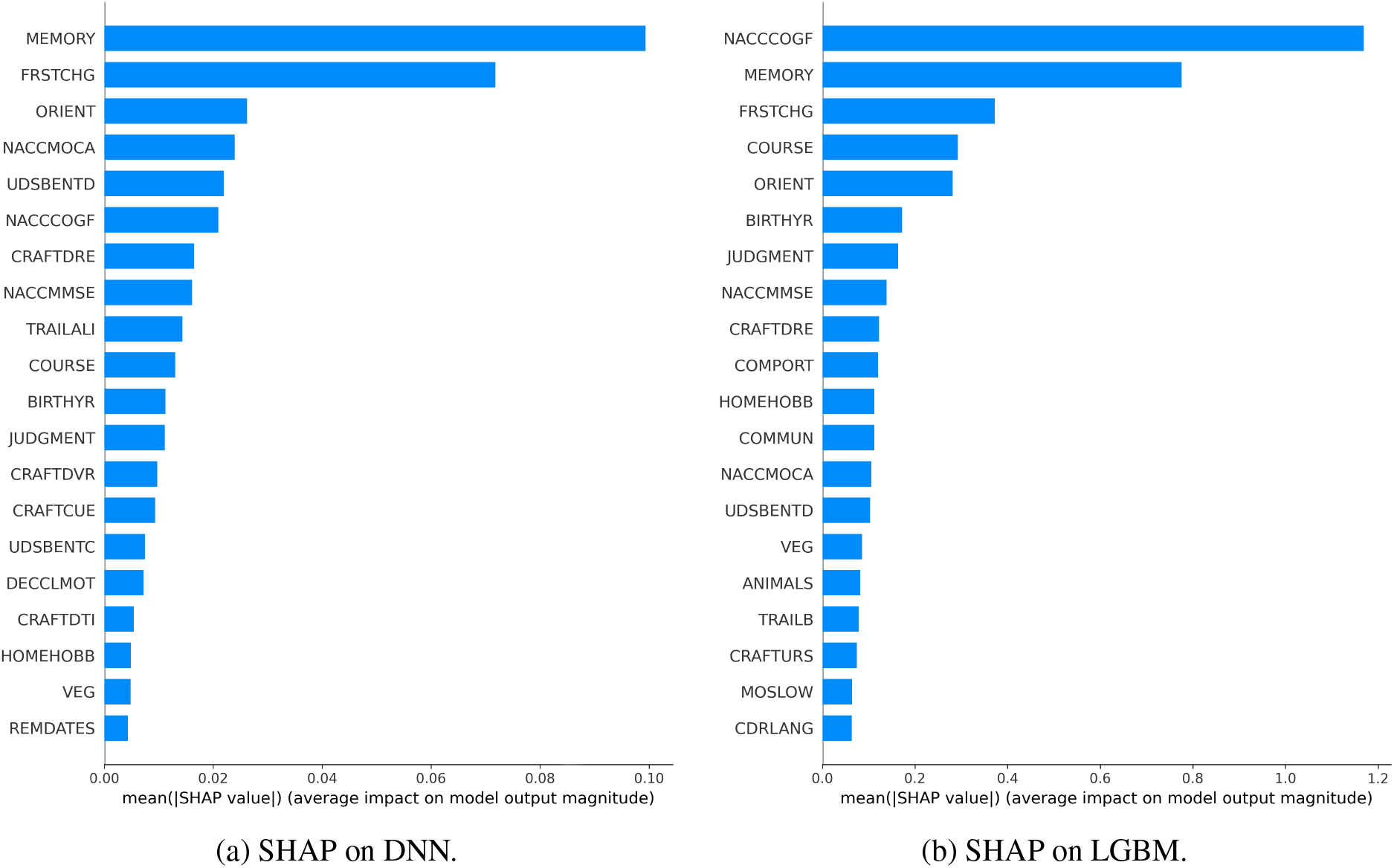
SHAP Global Explanations values.

The findings highlight the shared relevance of features such as MEMORY, NACCCOGF, FRSTCHG, ORIENT, and JUDGMENT, which consistently emerge as key predictors of Alzheimer’s disease (AD) across both DNN and LGBM models. Interestingly, while MEMORY remains the dominant predictor in both models, NACCCOGF exhibits substantially greater influence in LGBM compared to DNN, reflecting differences in how tree-based and neural network architectures allocate predictive importance across cognitive measures. These distinctions provide insight into model-specific feature sensitivities while confirming the central role of core cognitive and functional assessments in capturing early manifestations of AD, thereby enhancing the models’ predictive capacity.

Specifically, NACCCOGF reflects the predominant symptom initially recognized as a decline in cognition, often manifested in memory or executive dysfunction. The MEMORY domain further quantifies impairments in learning and recall. FRSTCHG identifies the cognitive domain where the earliest noticeable changes occurred, thus providing information on the initial trajectory of decline. Measures such as ORIENT (orientation to time, place, and person) and JUDGMENT (decision making and problem solving abilities) capture deficits in global cognition and executive function.

These results align with existing evidence that highlights the importance of symptoms and cognitive test scores as robust predictors of the risk of AD. A growing body of literature employing explainable AI techniques, particularly SHAP, has consistently identified measures related to memory, judgment, orientation, and communication-related measures as the core determinants of Alzheimer’s disease in various datasets and modeling approaches^13,34^.

#### Local Analysis: Individual Explanatory Power

To explore the local behavior of the model^35^, SHAP force plots were generated for a representative patient to visualize the contribution of each feature to the corresponding individual prediction. This type of visualization enables assessment not only of the relative importance of individual features, but also of the direction of their effect, indicating whether a given feature increases or decreases the predicted likelihood of an Alzheimer’s disease (AD) diagnosis.

In Figure 3, the deep neural network (DNN) explanation shows how the cumulative contribution of the displayed features shifts the model output toward an AD prediction at the selected visit. Figure 4 presents the corresponding local explanation for the same patient obtained using the LightGBM (LGBM) model. While both models identify overlapping cognitive and functional features as influential, differences in the magnitude and distribution of SHAP contributions reflect model-specific decision mechanisms.

**Figure 3:**
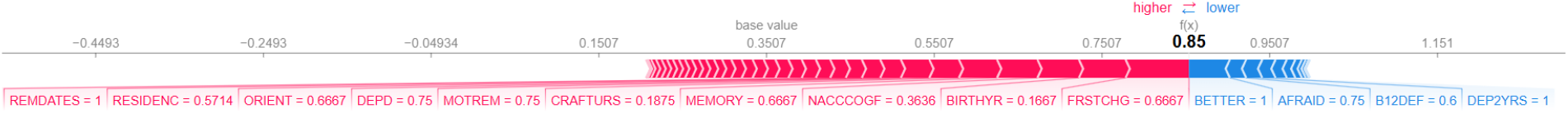
Individual-level SHAP force plot derived from the DNN model.

**Figure 4:**
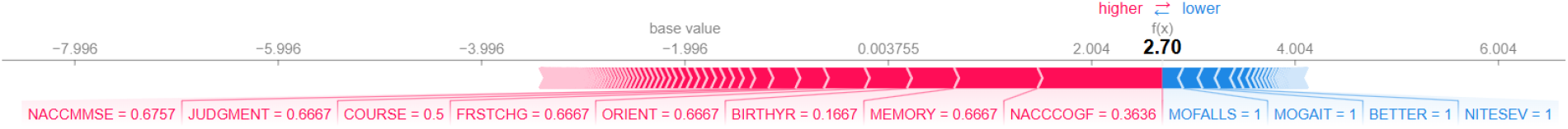
Individual-level SHAP force plot derived from the LGBM model for the same patient.

**Figure 5:**
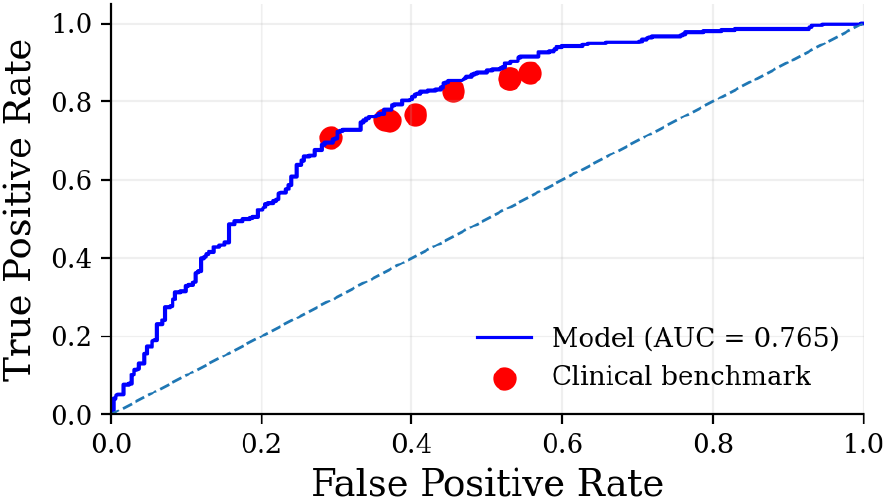
Comparison with clinicians accuracy

Such instance-level interpretability supports clinical validation of model outputs by facilitating the inspection of individual risk profiles and promoting the integration of AI-driven insights into diagnostic reasoning.

### 3.4 Comparison With Existing Literature

A rigorous benchmarking of the proposed framework requires contextualizing performance with respect to data constraints, feature selection strategies, and clinical applicability, rather than relying solely on absolute metrics. The following comparisons highlight how the proposed approach behaves under different methodological and clinical scenarios.

#### 3.4.1 Comparison with Standardized Cognitive Screening Tools

To further contextualize the efficacy of the proposed models, it is essential to compare their performance against established clinical screening tools, such as the Mini-Mental State Examination (MMSE)^36^ and the Montreal Cognitive Assessment (MoCA)^37^. These tests are widely implemented for rapid cognitive assessment in primary care; however, they often serve as single-point indicators rather than comprehensive predictive frameworks. In this comparison, we evaluated the diagnostic accuracy of these instruments by determining the optimal threshold for discriminating between AD and non-AD states. This was achieved by iteratively varying the classification cutoff for both NACCMMSE and NACCMOCA scores across their full standard range (0–30) with unit increments to identify the point of maximum accuracy; while the ROC-AUC was computed by considering the entire range of possible cutoffs to evaluate the overall discriminative capacity of each test.

The results, as reported in terms of both best-threshold Accuracy and overall ROC-AUC, underscore the limitations of using these tests as stand-alone predictors. Specifically, for the NACCMMSE, the optimal threshold was identified at 25.5, such that scores ≤ 25 indicate an AD classification, yielding an Accuracy of 0.803 and a ROC-AUC of 0.848. Similarly, for the NACCMOCA, the optimal threshold of 20.5, where scores ≤ 20 are classified as AD, resulted in an Accuracy of 0.813 and a ROC-AUC of 0.855. This performance demonstrates that while individual cognitive tests are robust indicators of impairment, the integration of multi-domain clinical data, the ones displayed in the NACC Categories in Table 1, allows our deep learning framework to capture more nuanced patterns of neurodegeneration, significantly exceeding the predictive power of traditional standalone screening tools.

#### 3.4.2 Comparison with lightweight models based on routine data

A first relevant comparison can be established with the work of Suresh et al. (2025)^12^, who evaluated both Artificial Neural Networks (ANN) and LightGBM models on the NACC dataset. A direct quantitative comparison is reported in Table 7, where the performance of both Artificial Neural Networks (ANN) and LightGBM models is evaluated on the NACC dataset. Notably, our proposed model consistently outperforms these baselines across all key metrics, demonstrating its superior predictive capability on similar selected feature.

**Table 7:** Performance comparison between models from Suresh et al. (2025) and the proposed approach on the NACC dataset. The first two rows report the results from Suresh et al., while the last two rows correspond to the models developed in this study.

| Model | Accuracy | Sensitivity | Specificity | Precision | F1-score | AUC |
| --- | --- | --- | --- | --- | --- | --- |
| ANN | 0.67 | 0.53 | 0.771 | 0.634 | 0.66 | 0.719 |
| LGBM | 0.82 | 0.780 | 0.795 | 0.836 | 0.79 | 0.91 |
| DNN | 0.895 | 0.877 | 0.905 | 0.833 | 0.854 | 0.961 |
| LGBM | 0.902 | 0.879 | 0.915 | 0.847 | 0.863 | 0.967 |

*Abbreviations:* AUC, Area Under the ROC Curve.

From the perspective of feature importance, a further distinction emerges between the two approaches. Both Suresh et al. and the present study employ SHAP-based analysis to interpret model predictions. However, their methodologies differ substantially in terms of the underlying feature space.

Suresh et al. apply SHAP to a highly reduced subset of 19 variables obtained through genetic algorithm–based feature selection, prioritizing model compactness. While this approach enables clear identification of the most influential predictors, it inherently constrains the analysis to a limited representation of the patient’s clinical profile.

In contrast, the present study performs SHAP analysis on a broader set of routinely collected clinical features, without enforcing aggressive dimensionality reduction. This results in a more distributed importance pattern, where multiple variables contribute meaningfully to the prediction. Notably, the most relevant features consistently map to core cognitive domains such as memory, orientation, and executive function, in agreement with established clinical understanding of Alzheimer’s disease^13^.

This difference suggests that, while both approaches achieve interpretability, the proposed methodology provides a more comprehensive view of the underlying decision process. By preserving a richer feature space, it becomes possible to capture the multifactorial nature of the disease more effectively, leading not only to improved predictive performance but also to interpretations that are more closely aligned with clinical reality.

#### 3.4.3 Comparison with unconstrained, performance-driven feature selection approaches

A second comparison concerns the work by Alatrany et al. (2024)^13^, which adopts a different methodological paradigm. In that study, the modeling pipeline begins with a high-dimensional feature space (over 1000 variables) and applies feature reduction techniques primarily aimed at maximizing predictive performance, without explicit constraints on data accessibility or acquisition cost.

In contrast, the present work imposes domain-driven constraints on feature selection, restricting the model to clinically accessible and scalable inputs. Despite this restriction, the resulting feature importance profiles show strong qualitative agreement with those identified in unconstrained approaches, particularly in relation to core cognitive domains such as memory, judgment, orientation, and communication .

This convergence suggests that the proposed framework is able to capture robust and clinically meaningful predictors, even under constrained conditions, thereby achieving a level of interpretability and consistency comparable to approaches optimized primarily for predictive performance.

#### 3.4.4 Comparison with historical clinical diagnostic accuracy (2005-2010)

A third comparison can be drawn with the study by Beach et al. (2012)^14^, which evaluates the accuracy of clinical diagnoses against neuropathological confirmation (gold standard) in the period 2005–2010. That study reports sensitivity values between approximately 70.9% and 87.3% and specificity between 44.3% and 70.8%, reflecting the limitations of clinical diagnosis prior to the widespread integration of advanced imaging techniques.

To ensure a meaningful comparison, we restrict our analysis to a subset of data aligned with gold-standard confirmation (PET-based), which introduces a more stringent evaluation setting. Under these conditions, the proposed model achieves an AUC of approximately 0.765.

Although this value is lower than the performance obtained on the full dataset, it remains comparable to the diagnostic accuracy of clinicians operating in pre-imaging clinical workflows, where similar information constraints applied. Therefore, in settings characterized by limited access to advanced biomarkers, the proposed approach can be considered competitive with historical clinical practice, while offering advantages in standardization, scalability, and reproducibility.

## 4 Conclusion

This study demonstrates that interpretable deep learning framework can effectively model a presumptive etiological diagnosis of Alzheimer’s disease. By learning from diagnostic labels that encode expert clinical judgment, rather than neuropathological ground truth, the proposed approach captures clinically meaningful decision patterns, achieving high predictive performance while remaining aligned with the practical constraints of routine diagnosis.

Through rigorous pre-processing, feature selection, and hyperparameter optimization, both DNN and LGBM achieved an AUC > 0.96 and an accuracy of almost 90%, demonstrating robust discrimination between AD and non-AD cognitive states.

Crucially, SHAP-based global and local interpretability analyses revealed feature attributions consistent with established clinical knowledge, supporting the internal validity of the learned representations. This interpretability is particularly relevant in the absence of neuropathological ground truth, as it enables verification that the model’s predictions are grounded in clinically meaningful patterns rather than spurious correlations.

These findings highlight the feasibility of deploying explainable machine learning tools for scalable and cost-effective early detection of Alzheimer’s disease, particularly in primary care settings and resource-limited environments, or remote screening scenarios. While not intended to replace definitive diagnostic procedures, such tools may facilitate early risk stratification and timely referral for advanced evaluation.

Future research will leverage the longitudinal NACC dataset to model patient-specific disease trajectories, transitioning from static diagnostics to personalized prognostic modelling. Integrating temporal deep learning architectures, such as RNNs and LSTMs, with survival analysis methods, alongside strategies for handling missing longitudinal data, will improve forecasting of progression and time-to-event outcomes. Furthermore, developing a multiclass classification framework to distinguish stages of cognitive decline using only tabular clinical and neuropsychological data could improve patient stratification, supporting timely, personalized, and targeted therapeutic interventions.

We conclude by emphasizing that the present work should be interpreted as a preliminary feasibility study aimed at evaluating whether explainable machine learning models trained exclusively on routine and low-cost clinical data can achieve clinically meaningful discrimination of Alzheimer’s disease. In this perspective, future developments of the project will focus not only on longitudinal model refinement through continuous data acquisition, but also on translational validation in collaboration with hospitals and specialized neurological research centres. The low-cost, non-invasive, and highly scalable nature of the proposed framework makes it particularly suitable for integration into real-world clinical pathways, telemedicine infrastructures, and preliminary screening programs, where rapid identification of high-risk individuals could support clinicians in prioritizing referrals for advanced diagnostic assessment.

## Data Availability

All data produced in the present study are available upon reasonable request to the authors.

## Data Availability

The NACC Dataset is openly available for researchers at https://naccdata.org/with a data access request.

## Acknowledgements

We would like to thank The National Alzheimer’s Disease Coordinating Center (NACC) for access to NACC data.

The NACC database is funded by NIA/NIH Grant U24 AG072122. NACC data are contributed by the NIA-funded ADRCs: P30 AG062429 (PI James Brewer, MD, PhD), P30 AG066468 (PI Oscar Lopez, MD), P30 AG062421 (PI Bradley Hyman, MD, PhD), P30 AG066509 (PI Thomas Grabowski, MD), P30 AG066514 (PI Mary Sano, PhD), P30 AG066530 (PI Helena Chui, MD), P30 AG066507 (PI Marilyn Albert, PhD), P30 AG066444 (PI David Holtzman, MD), P30 AG066518 (PI Lisa Silbert, MD, MCR), P30 AG066512 (PI Thomas Wisniewski, MD), P30 AG066462 (PI Scott Small, MD), P30 AG072979 (PI David Wolk, MD), P30 AG072972 (PI Charles DeCarli, MD), P30 AG072976 (PI Andrew Saykin, PsyD), P30 AG072975 (PI Julie A. Schneider, MD, MS), P30 AG072978 (PI Ann McKee, MD), P30 AG072977 (PI Robert Vassar, PhD), P30 AG066519 (PI Frank LaFerla, PhD), P30 AG062677 (PI Ronald Petersen, MD, PhD), P30 AG079280 (PI Jessica Langbaum, PhD), P30 AG062422 (PI Gil Rabinovici, MD), P30 AG066511 (PI Allan Levey, MD, PhD), P30 AG072946 (PI Linda Van Eldik, PhD), P30 AG062715 (PI Sanjay Asthana, MD, FRCP), P30 AG072973 (PI Russell Swerdlow, MD), P30 AG066506 (PI Glenn Smith, PhD, ABPP), P30 AG066508 (PI Stephen Strittmatter, MD, PhD), P30 AG066515 (PI Victor Henderson, MD, MS), P30 AG072947 (PI Suzanne Craft, PhD), P30 AG072931 (PI Henry Paulson, MD, PhD), P30 AG066546 (PI Sudha Seshadri, MD), P30 AG086401 (PI Erik Roberson, MD, PhD), P30 AG086404 (PI Gary Rosenberg, MD), P20 AG068082 (PI Angela Jefferson, PhD), P30 AG072958 (PI Heather Whitson, MD), P30 AG072959 (PI James Leverenz, MD).

## Author contributions

Daniele De Carli and Alberto Sudati contributed equally to all aspects of this work, including study design, data analysis, model development, and manuscript preparation. Fabio Dercole supervised the research and contributed to the critical revision of the manuscript.

